# Food Insecurity and High-Risk Sexual Behaviour among Women of Reproductive Age in Zambia: A Socio-Ecological Model

**DOI:** 10.64898/2026.02.10.26345965

**Authors:** Elizabeth T. Nyirenda, Clifford O. Odimegwu, Latifat Ibisomi

**Author notes:** Corresponding author: Elizabeth T. Nyirenda.

## Abstract

High-risk sexual behaviour has been shown to be influenced by interpersonal, social, and economic factors in different situations and been linked to food insecurity. This study therefore investigated the individual, household and community factors associated with high-risk sexual behaviour among women age 18-49 in Zambia, paying particular attention to food insecurity status. It was anchored on the Socio-Ecological Model and used a cross-sectional non-intervention study design. We fitted survey weighted logistic regression models based on 1784 weighted cases to examine the association between food insecurity and high-risk sexual behaviour and determine the predictors of high-risk sexual behaviour.

The results show that 44 percent of the women engaged in of high-risk sexual. We found that 53.9 (±6.3) percent of the women age 18-49 experienced moderate to severe food insecurity while 21.2 (±2.9) percent experienced severe food insecurity. Food insecurity is associated with high-risk sexual behaviour before controlling for other covariates. Our results show that women with higher education [aOR= 0.471 95%CI 0.254,0.872], women who were married [aOR= 0.123 95%CI 0.065, 0.231], women whose most recent pregnancy was intended [aOR= 0.482 95%CI 0.328, 0.709], women in the rich wealth tertile [aOR= 0.372 95%CI 0.188, 0.736] and women in rural areas[aOR= 0.425 95%CI 0.239, 0.758] had lower odds of engaging in high-risk sexual behaviour. Our findings confirm that high-risk sexual behaviour is influenced by factors beyond the individual level. Thus, HIV prevention response should be holistic in nature. Greater attention should be paid to adolescent girls to reduce high-risk sexual behaviour and avert possible consequences. Improving the social-economic status of women through education and economic empowerment among other interventions, will advertently reduce vulnerability to HIV infection, unintended pregnancy, improve the health and well-being of women and ensure that women are not left behind.

## Introduction

High-risk sexual behaviour is defined as any sexual activity that increases the risk of acquiring sexually transmitted infections and unwanted pregnancies [1]. High-risk sexual behaviour has been shown to be influenced by interpersonal, social, and economic factors in different situations [2]. High-risk sexual behaviour has also been linked to food insecurity [3, 4]. Food insecurity exists when the availability of nutritionally adequate and safe foods or the ability to acquire acceptable foods in socially acceptable ways is limited or uncertain [5].

High-risk sexual behaviours have increased the burden of STIs, HIV and unintended pregnancies globally. It is estimated that more than 1 million STIs are acquired every day. In 2020, WHO estimated 374 million new infections every year, including 129 million cases of chlamydia, 82 million cases of gonorrhoea, 7.1 million cases of syphilis and 156 million cases of trichomoniasis [6]. An estimated 40.8 million people globally were living with HIV and 1.5 million people became newly infected with HIV in 2024 [7]. In addition, nearly half of all pregnancies, totalling 121 million each year throughout the world, are unintended [8]. Further, about 29.6 percent of the global population (2.4 billion people) were moderately or severely food insecure in 2022, 391 million more people than in 2019. Food insecurity affects more women than men in every region of the world [9].

Zambia is among the countries with highest burden of HIV [10] compounded with high-risk sexual behaviours which have worsened over the years [11]. HIV prevalence remains high at 11.0% among adults 15 years and older. With higher prevalence among women (13.9%) compared with men (8.0%) [10]. The number of new HIV cases remains high especially among women. HIV incidence is estimated at 28,000 new cases of HIV per year among adults 15 years and older, with higher incidence among women (0.56%) compared to men (0.06%) [10]. Zambia is also faced with increasing food insecurity especially considering changes in rainfall patterns and macroeconomic instability [12].

Literature shows that women are the most affected by food insecurity and may resort to high-risk sexual behaviour to provide for themselves and their families [13–15]. Hunger can increase vulnerability to HIV by increasing risk taking behaviour [15]. We therefore hypothesise that food insecurity is likely to increase due to drought and in turn increase high-risk sexual behaviour among women in Zambia. Increase in high-risk sexual behavior has the potential to negate gains made in HIV response and threaten the goal of ending AIDS as a public health concern by 2030 [16]

Previous research has focused on people living with HIV and the effect of food insecurity on adherence to treatment [13, 14, 17, 18]. Few studies have investigated the association of food insecurity and high-risk sexual behaviour among women of reproductive age and examined covariates beyond the individual level. Thus, the role of food insecurity on high-risk sexual behaviour among women of reproductive age remains largely understudied. This study, therefore, investigates the individual, household and community level factors associated with high-risk sexual behaviour among women of reproductive age in Zambia, paying particular attention to food insecurity status. This paper includes the materials and methods used and the findings of the study. We also discuss the findings of our study in relation to the findings of previous studies.

## Materials and Methods

The study utilized primary data collected from 1787 sexually active women age 18-49 who had lived in the study sites for at least 12 months. A structured questionnaire was used to interview women aged 18-49 who consented to participate in the survey from their households. We collected information on household members, household characteristics, assets and possession, food insecurity experience, sexual activity, contraception, pregnancy and reproduction and cultural norms and social support. A computer aided personal interviewing (CAPI) application developed using Census and Survey Processing System (CSPro) version 7.7.2- a public domain software that supports data collection and transfer using android devices was used for data collection.

Two provinces were purposively selected for the study. Lusaka province was selected as the urban province and Western province was selected as the rural province. Lusaka province has the highest HIV prevalence of 14.4 % in the population 15 years and older. It also has the second highest percentage (34.8%) of women reporting high-risk sex (sexual intercourse with a non-marital, non-cohabiting partner) in the 12 months prior to the 2021 ZAMPHIA survey. It also had the highest HIV prevalence among women 15 years and older at 18.6% [10]. Western province has the second highest HIV prevalence in the population 15 years and older at 13.6%, the highest percentage of women age 15-24 (27.9%) who reported having sexual intercourse before age 15 and the highest percentage of women reporting high-risk sex at 39.4% in the last 12 months prior to the 2021 ZAMPHIA survey [10].

Two districts, one rural (Kaoma) and one urban (Lusaka) were identified to be the sample sites where the survey would be undertaken. The sample in the two districts, Kaoma and Lusaka, was selected in multiple stages. In the first stage, four wards were selected in each district. In the second stage, eight (8) Enumeration Areas were selected in each of the wards using Probability Proportional to size (PPS). The measure of size was the number of households captured in the EAs during the 2022 Census of Population and Housing.

We used the formula for stratified random sampling proposed by Lemeshow et al. [19] to determine the sample. The estimation formula indicated below was used to determine the sample.

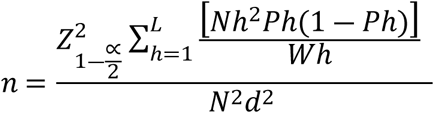

Using equal allocation in each stratum, assuming a 95% confidence interval and a precision of ±3 percent and power of >90%. The required sample was 1843 women. To achieve the target sample, an average of forty (40) households were selected and interviewed in each cluster. In each household, one-woman age 18-49 years was selected and interviewed. A total of 1803 interviews were conducted resulting in a 97.8 percent response rate. Out of the 1803 interviews, 943 interviews were conducted in Lusaka while 860 were conducted in Kaoma. To ensure that the sample is representative of the two districts, the data were weighted.

### Variables

The primary outcome of interest in this study is high-risk sexual behaviour. It is a dichotomous variable coded 0 “No” if a woman did not report any high-risk sexual behaviour and 1 “Yes” if a woman reported at least one high-risk sexual behaviour. It is constructed using five risky sexual behaviours. These are: (1) condom non-use with nonmarital or cohabiting partner, defined as condom non-use during sex with a non-marital or non-cohabiting partner [20]; (2) multiple sexual partners, defined as having two or more sexual partners [21]; (3) early sexual debut, defined as having sexual intercourse before age 15 [22]; (4) transactional sex, defined as ever engaged in sexual intercourse in exchange for money, food, gift or other material benefit [23]; (5) intergenerational sex, defined as having a sexual partner/s who is 10 or more years older [24].

#### Main Predictor Variable

The primary explanatory variable of interest is household food insecurity. This variable was measured using Food Insecurity Experience Scale Survey Module (FIES-SM). The FIES-SM is a validated measure developed by the Food Agriculture Organisation to estimate food insecurity prevalence. It is composed of eight core questions with dichotomous yes/no responses [25]. The FIES measures access to food at household or individual level. For this study, food insecurity was measured at household level. We used the 12-month household referenced FIES-SM to estimate household food insecurity in the last 12 months prior to the survey (12 months prior to October 2024).

Women age 18-49 were asked if, during the last 12 months, there was a time when, because of lack of money or other resources: they were worried they would not have enough food to eat; they were unable to eat healthy and nutritious food; they ate only a few kinds of food; they had to skip a meal; they ate less than they thought they should; their household ran out of food; they were hungry but did not eat and if they went without eating for a whole day [25].

We measured food insecurity in two ways. The first measure is an interval measure based on the global standards threshholds. The global standards thresholds was used to estimate moderate to severe food insecurity and severe food insecurity. The threshold for moderate to severe food insecurity is −0.31 set at the FIES item severity level ATELESS and the threshold for severe food insecurity is 1.88 set at the FIES item severity level WHLDAY. The second measure is an ordinal measure based on discrete assignment of raw scores. A raw score of “0” was categorized as food secure, a raw score of 1-3 was categorized as mild food insecurity, a raw score of 4-6 was categorized as moderate food insecurity and a raw score of 7 or 8 was categorized as severe food insecurity [26].

##### Other Predictor Variables

Individual level predictors included age (Coded 1 if 18-19, 2 if 20-24, 3 if 25-29, 4 if 30-34, 5 if 35-39, 6 if 40-44 and 7 if 45-49), education (Coded 0 if no education, 1 if primary, 2 if secondary, 3 if higher), marital status (Coded 0 if Never married, 1 if Married/ Living together and 3 if Divorced/ Widowed/ Separated),working status (Coded 0 if not working and 1 if working), parity (Coded 1 Nulliparous (no children) and 2 if Primiparous or Multiparous (one or more children), contraceptive use (Coded 0 if No and 1 if Yes), comprehensive knowledge of HIV(Coded 0 if respondent has no comprehensive knowledge of HIV and 1 if respondent correctly identifies the two major ways of preventing sexual transmission of HIV and rejects the two most common local misconceptions and know that a healthy-looking person can have HIV) [20], bodily autonomy (Coded 1 “Low autonomy” if score is 1 or 2, and 2 “Average autonomy” if score is 3, and, 3 “High autonomy” if score is 4 or 5), use of pre-exposure prophylaxis (Coded 0 if No and 1 if Yes), pregnancy intendedness (Coded 0 if unintended, 1 if intended), test for HIV(Coded 0 if No and 1 if Yes), away from home for more than a month in the last 12 months (Coded 0 if No and 1 if Yes), sexually transmitted infection in the last 12 months (Coded 0 if No and 1 if Yes) and age at first birth (Code 1 if below age 20 and 2 if above age 20).

Household level variables included wealth index (Coded 1 Poor 2 if Middle and 3 Rich) [27], household size (Coded 1 Small if household size is 1-3, 2 Medium if household size is 4-6 and, 3 Large if household size is 7 and over) [28] sex of household head (Coded 1 if Male, 2 if Female) and living arrangements (Coded 1 “Lives alone”, 2 “Lives with own family”, and, 3 “Lives with others”) [29].

Community level variables included family support (Coded 0 if No and 1 if Yes, place of residence (Coded 1 if Urban and 2 if Rural), media exposure (Coded 0 if No and 1 if Yes), sexuality education (Coded 0 if No and 1 if Yes), social support (Coded 0 if No and 1 if Yes) and cultural norms (Coded 0 if No and 1 if Yes).

### Statistical Analysis

Prior to estimation of food insecurity prevalence rates, we subjected our FIES data to statistical validation using the Rasch model. Using the FAO ShinyApp [30], we computed item statistics (Infit and Outfit) based on 704 complete and non-extreme scores. For statistical validation, cases between 300 and 999 provide item severity parameters that are quite reliable. An adequate fit of the Rasch model is indicated by infit and outfit statistics of 0.7-1.3 for each item. Infit of greater than 1.3 and outfit greater than 2 are considered high. Items with high infit should be dropped from the analysis. We kept all 8 FIES items in the analysis as infit and outfit was within acceptable ranges [25].

We also considered the Rasch reliability to provide information about the discriminatory power of the overall scale, measuring the proportion of variability in the data that is explained by the Rasch model. For an 8-item FIES scale, a Rasch reliability value above 0.7 is considered acceptable [25]. Our analysis shows a Rasch reliability of 0.734014. We also employed automatic equating (based on the minimum correlation plot) with the minimum correlation set at 0.97 to plot the number of common and unique items and to compute the percentage of correlation in the FIES items. For sound estimation, at least 5 items should be common. Based on the results of the infit, outfit, Rasch reliability and equating we are confident that our data is of good quality for estimation of food insecurity.

We used STATA version 14.2 for data wrangling and analysis. Analysis of high risk-sexual behaviour was done at three levels: univariate, bivariate and multivariate. Univariate analysis was used to analyse the characteristics of respondents and estimate the prevalence of high-risk sexual behaviours. Bivariate analysis was used to examine the association between high-risk sexual behaviour and individual, household and community variables. Chi-squared analysis was used to test association between the predictor variables and high-risk sexual behaviour (P<0.05). We fitted logit models to further examine the association between high-risk sexual behaviour and selected predictor variables.

We report unadjusted odds ratios for associations between high-risk sexual behaviour and selected variables significant at P<0.05. We also report adjusted odds ratios for associations between high-risk sexual behaviour and selected variables significant at P<0.05. Collinearity diagnostics for the fitted logit models were performed. All covariates included in the model have a VIF less than 5 indicating no problem of multicollinearity [31, 32]. All data wrangling and analysis was done using Stata version 14.2 except for statistical validation of FIES data and estimation of food insecurity prevalence rates which was done using the FAO ShinnyApp.

### Ethics approval and consent to participate

The study obtained ethical approval from a Zambian ethical review board (ERES Converge) and ethical approval from the University of the Witwatersrand. The study also obtained approval of authority to conduct research from the National Health Research Authority of Zambia and obtained a letter of permission to conduct research from the Ministry of Health.

The study was approved by the University of the Witwatersrand, Faculty of Humanities and the University of Witwatersrand Human Research Ethics Committee (Non-medical) Protocol number H24/06/34. The principal investigator was also certified in research ethics by the University of the Witwatersrand and obtained certification as a health Researcher from the Zambia National Health Research Authority.

All research instruments used were approved, pretested and administered by trained research assistants. All instruments, information sheets and consent forms were translated into Lozi and Nyanja-the local languages used in the study sites.

Persons eligible to participate in the study were provided with an information sheet, after comprehending the information, they voluntarily consented to participate in the study. The information sheet provided to participants contained the study aims, methods, potential risks, anticipated benefits and right to withdraw. Research Assistants ensured a safe space to administer the questionnaire to ensure confidentiality and avoid compromising the safety of study participants.

All consent was obtained in written. Participants who were unable to write were provided with ink. The study observed the ethical principles of: respect for persons, beneficence and non-maleficence and justice. The data collected and the consent forms have been archived securely and will not be shared with any unauthorised persons. In the event of distress during the data collection process, a distress protocol was specified.

## Results

The study analysed 1784 weighted cases of sexually active women age 18-49 to examine the individual, household and community level factors associated with high-risk sexual behaviour, paying particular attention to food insecurity status. Even though 1803 women participated in the survey, only 1787 had complete FIES data. Thus, the final analysis is based on 1784 weighted cases.

### Individual, household and community level characteristics of study participants

The study collected information on background characteristics of women, household characteristics, food insecurity experience, sexual activity, contraception, pregnancy and reproduction and cultural norms and social support. Table 1 shows the individual characteristics of study participants. The results show that about 25% of participants were food secure and 30% experienced severe food insecurity (based on discrete assignment of raw scores). The 20-24 age group had the highest percentage of participants followed by the 25-29 age group at 22%. The 18-19 age group had the lowest percentage of participants at 7%. The average age of participants was 30.4 years and the median age was 29.0 years. The results show that 54% of the participants had secondary level education, 27% had primary level education and 18% reported having higher level of education. The results also show that 46% of the participants were married, 39% were never married and 16% were widowed, separated or divorced. Further, 53% of the participants reported that they do some work to earn an income. Regarding parity, 68% of the participants were primiparous/multiparous. About 48% of participants reported that they were currently using a method of contraception to delay or avoid getting pregnant. In addition, 80% of participants had comprehensive knowledge of HIV and about 10% reported having used pre-exposure prophylaxis. Forty-Eight percent of the participants reported that their last pregnancy was unintended and 39% reported having their first birth below age 20.

**Table 1:** Individual characteristics of study participants.

The survey collected data on household members, possessions and characteristics. Table 2 shows the household level characteristics of study participants. The results show that 53% of the participants were in the rich wealth category, 38% were in the middle wealth category while 9% were in the poor wealth category. Forty-nine percent of the participants resided in households that had a small household size (1-3 members) while only 9% reported residing in households with a large household size (7+ members). The results also show that 56% of the participants were from male headed households while 44% were from female headed households. Further, 79% of the participants were living with their families, 12 % lived alone and 9% were living with others.

**Table 2:** Household characteristics of study participants.

The survey also collected data on community level characteristics. Table 3 shows the community level characteristics of study participants. The results show that 37% of the participants indicated receiving support from family members. About 80% reported not having any form of sexuality education in their communities. Eighty-eight percent were exposed to media (Radio, TV or Internet) at least once a week. Ninety-one percent of the women indicated being affected by drought. About 88% and 86% of the women rated the cost of living and food prices as high, respectively. In addition, 19% indicated having cultural norms supporting high-risk sexual behaviour in their community.

**Table 3:** Community level characteristics of study participants.

#### Prevalence of high-risk sexual behaviour

We constructed a dichotomous composite high-risk sexual variable based on the five high-risk sexual behaviour variables – Condom non-use, Multiple sexual partnerships, Early sexual debut, Transactional sex and Intergenerational sex, coded “Yes” for women who engaged in any of the five high-risk behaviours and “No” for women who did not indicate engaging in any of the five behaviours. Figure 1 shows the prevalence of high-risk sexual behaviour and each of the five high-risk sexual behaviours.

**Figure 1:** Prevalence of high-risk sexual behavior.

The results show that 44 percent of the women age 18-49 engaged in high-risk sexual behaviour. Disaggregation by five high-risk sexual behaviours shows that 47% of women who indicated having sex with a non-marital or non-cohabiting partner did not use a condom at last sex. Six percent reported having more than one sexual partner in the last 12 months prior to the survey. Seven percent reported having intercourse before the age of 15. About 6 percent reported engaging in transactional sex and 17 percent had a partner who is 10 years or more older than them.

### Prevalence of food insecurity

The 8 item,12 months referenced household FIES-SM was used to estimate the prevalence of moderate to severe food insecurity and severe food insecurity. The results show that 53.9 (±6.3) percent of the women age 18-49 experienced moderate to severe food insecurity while 21.2 (±2.9) percent experienced severe food insecurity. The results also show that food insecurity is higher in rural areas compared with urban areas (Figure 2)

**Figure 2:** Prevalence of food insecurity.

This study investigated the association between food insecurity and high-risk sexual behaviour. Thus, we disaggregated the prevalence of food insecurity by high-risk sexual behaviour. Table 4 shows the prevalence of food insecurity by high-risk sexual behaviour. The results show that moderate to severe food insecurity and severe food insecurity is higher among women 18-49 years who reported engaging in high-risk sexual behaviour across all high-risk sexual behaviour variables including the composite high-risk sexual behaviour variable. Intergenerational sex shows slightly lower severe food insecurity among women who reported engaging in intergenerational sex, however, the margin of error indicates higher prevalence among this group of women compared to their counterparts who did not report engaging in intergenerational sex.

**Table 4:** Prevalence of food insecurity by high-risk sexual behaviour.

Table 5 shows a summary of findings of chi-squared analysis comparing the variation of individual, household and community level variables with composite high-risk sexual behaviour. The results show that food insecurity, age, education, marital status, working status, use of pre-exposure prophylaxis, pregnancy intendedness, had STI in the last 12 months prior to the survey, age at first birth, wealth index, sex of household head and place of residence were significantly associated with high-risk sexual behaviour (P<0.05)

**Table 5:** Prevalence of high-risk sexual behaviour by selected individual, household, community level variables.

Table 6 shows the results of the survey-weighted unadjusted and adjusted odds of high-risk sexual behaviour and selected individual, household and community level variables. The Unadjusted odds show that high-risk sexual behaviour is associated with moderate and severe food insecurity. Women who were moderately food insecure had higher odds of engaging in high-risk sexual behaviour compared with women who were food secure [ uOR= 1.606 95%CI 1.038, 2.486]. Women with severe food insecurity had even higher odds [uOR= 1.826 95% CI 1.321, 2.525] of engaging in high-risk sexual behaviour compared with women who were food secure.

**Table 6:** Unadjusted and adjusted associations between High-risk sexual behaviour and selected individual, household and community level variables.

The unadjusted odds also shows that women age 25-29 had lower odds of engaging in high-risk sexual behaviour compared with women age 18-19 [uOR= 0.582 95%CI 0.374,0.905]. Women age 30-34 had significantly lower odds of engaging in high-risk sexual behaviour compared with women age 18-19 [uOR= 0.491 95%CI 0.305, 0.793].

The unadjusted odds also show that women with higher level of education, had significantly lower odds of high-risk sexual behaviour compared with women with primary level of education [uOR= 0.426 95%CI 0.287,0.633]. Women who were married had lower odds of engaging in high-risk sexual behaviour compared with women who were never married [uOR = 0.283 95%CI 0.200,0.401]. Women who were widowed, separated or divorced had higher odds of engaging in high-risk sexual behaviour compared with never married women [uOR = 2.191 95% CI 1.282,3.746]. Women who were working had higher odds of high-risk sexual behaviour compared with those who were not working [uOR = 1.627 95% CI 1.167, 2.270]. Women who reported ever using pre-exposure prophylaxis had higher odds of engaging in high-risk sexual behaviour compared to those who reported not ever using pre-exposure prophylaxis [uOR = 2.218 95% CI 1.536, 3.204].

Further, women whose most recent pregnancy was intended had lower odds of engaging in high-risk sexual behaviour compared with women who reported that their most recent pregnancy was unintended [uOR = 0.487 95%CI 0.347, 0.682]. Women who reported having an STI in the 12 months prior to the survey had higher odds of engaging in high-risk sexual behaviour compared with women who reported not having any STI in the 12 months prior to the survey [uOR = 1.925 95% CI 1.188,3.120]. Women who had a first birth after age 20 had lower odds of engaging in high-risk sexual behaviour compared with women who had their first birth before age 20 [ uOR = 0.615 95%CI 0.486, 0.777].

In addition, women in the rich wealth tertile, had lower odds of engaging in high-risk sexual behaviour compared with women in the poor wealth tertile [uOR = 0.531 95% CI 0.366,0.770]. Women in female headed households had higher odds of high-risk sexual behaviour compared to women in male headed households [uOR= 2.925 95% CI 2.378, 3.598].

Table 6 also shows the adjusted associations between high-risk sexual behaviour and selected individual, household and community variables. The adjusted odds show that food insecurity is not associated with high-risk sexual behaviour after controlling for other covariates. The results show that women age 20-24 [aOR= 0.319 95% CI 0.112,0.903] and women age 30-34 [aOR = 0.269 95% CI 0.740,0.976] have lower odds of high-risk sexual behaviour compared to women age 18-19.

The results also show that women with higher education level have lower odds of engaging in high-risk sexual behaviour compared to women with primary education level [aOR= 0.471 95%CI 0.254,0.872]. Further, the results show that women who were married had lower odds of engaging in high-risk sexual behaviour compared with never married women [aOR = 0.123 95%CI 0.065, 0.231]. Women whose most recent pregnancy was intended had lower odds of high-risk sexual behaviour compared with women whose most recent pregnancy was unintended [aOR = 0.482 95%CI 0.328, 0.709].

Regarding wealth quintile, women in the rich wealth quintile, have lower odds of engaging in high-risk sexual behaviour compared with women in the poor wealth quintile [aOR = 0.372 95%CI 0.188,0.736]. In addition, women in rural areas have lower odds of high-risk sexual behaviour compared to women in urban areas [aOR= 0.425 95%CI 0.239, 0.758].

## Discussion

The study set out to measure the level of food insecurity and high-risk sexual behaviour, and examine the predictors of high-risk sexual behaviour among women age 18-49 in Zambia paying particular attention to food insecurity status. The results show that moderate to severe food insecurity was 53.9% (±6.3) and severe food insecurity was estimated at 21.2% (±2.9). This indicates a high level of food insecurity among women age 18-49. Data collection for this study was undertaken from October 6-29, 2024. On 29 February 2024, the Zambian Government declared a state of disaster in response to an El Niño-induced drought during the 2023–2024 agricultural season. The drought significantly affected agriculture and electricity production, heightening food insecurity and affecting livelihoods in 84 out of 116 districts [12]. Thus, the high estimates of moderate to severe and severe food insecurity among women could be explained by the drought situation.

We also found that 47 percent of women who reported having intercourse with a non-marital or non-cohabiting partner did not use a condom at last sexual intercourse in the 12 months prior to the survey. Six percent of the women indicated having more than one sexual partner. Seven percent indicated having sex before the age of 15 and 6 percent reported having sex in exchange for goods, money or other benefits. Seventeen percent indicated having a sexual partner who is 10 years or older than them.

We estimated high-risk sexual behaviour at 44 percent based on five high risk sexual behaviours investigated in this study - condom non-use, multiple sexual partnerships, early sexual debut, transactional sex and intergenerational sex. Overall, the percentage of women who engage in behaviour that put them at risk of acquiring HIV and intended pregnancy is high. The 2021 ZAMPHIA survey indicated that HIV incidence is higher among women compared to men in Zambia. High-risk sexual behaviour is a persistent driver of HIV infections [33, 34]. Thus, HIV incidence among women will continue to be high if high-risk sexual behaviours persist.

We fitted a survey weighted logistic regression models to examine the predictors of high-risk sexual behaviour paying particular attention to food insecurity status. Anchored on the socioecological model, this study examined the individual, household and community level predictors associated with high-risk sexual behaviour. The unadjusted odds shows that food insecurity, age, education, marital status, working status, use of pre-exposure prophylaxis, pregnancy intendedness, having an STI in the 12 months preceding the survey, age at first birth, wealth index, sex of household head and residence are associated with high-risk sexual behaviour.

We found that food insecurity was not significantly associated with high-risk sexual behaviour after controlling for other covariates, however, the adjusted odds ratios show a direct relationship between food insecurity and high-risk sex behaviour for women who experienced moderate food insecurity and those who experienced severe food insecurity. The results of the adjusted model show that age, education, marital status, pregnancy intendedness, wealth index and residence were associated with high-risk sexual behaviour. Women age 20-24 and 30-34 had lower odds of engaging in high-risk sexual behaviour. Women with higher education, those who were married/ living together with a partner, women whose last pregnancy was intended, women who were in the rich wealth tertile and women in rural areas had lower odds of engaging in high-risk sexual behaviour.

We found that adolescents 18-19 had higher odds of engaging in high-risk sexual behaviour compared to women who were older. Uchidi also found that younger women are significantly more likely to report high-risk sexual behaviour than older women [35]. Our findings reflect the high number of new HIV infections and teenage pregnancy observed among adolescents in Zambia [10, 11]. According to Leclerc-Madlala, without a developmental approach that can address and dismantle the cultural legacies (e.g. cultural scripts that allow the intertwining of sex and material giving) that tend to affirm and lend legitimacy to cross-generational and multiple partnerships, it is not realistic to expect young women to forgo in a near future the many potential benefits that come from involvement in high-risk sexual behaviour [36].

Our results show that marriage is protective against high-risk sexual behaviour among women of reproductive age. Our results align with the findings of Arabi-Mianrood et.al [37] and also congruent with Uchidi et. al [35] who found that marriage was protective against high-risk sexual behaviour among women.

We also found that women with higher education have lower odds of high-risk sexual behaviour. Previous studies confirm that education is protective against high-risk sexual behaviour [35, 38–40].

Our results also show that women whose most recent pregnancy was intended had lower odds of high-risk sexual behaviour. This finding indicates that women with unintended pregnancy may be engaging in high-risk sexual behaviour. It also suggests that women with intended pregnancy are in stable unions which are protective against high-risk sexual behaviour.

We also found that women from rich wealth index had lower odds of engaging in high-risk sexual behaviour. Our findings are congruent with the findings of Uthman, Puplampu et al., and Workneh et.al [40–42].

We hypothesised that women in rural areas are more likely to engage in high-risk sexual behaviour due to heightened vulnerability due to increased food insecurity resulting from the drought. Women in rural areas face increased risk due to increased dependence on the environment compared with urban women. Interestingly, our results show that women in rural areas have lower odds of engaging in high-risk sexual behaviour compared to women in urban areas. Our findings are not congruent with the findings of previous studies that found high- risk sexual behaviour to be more prevalent in rural areas [35, 40]. It should be noted that this study collected data from Lusaka and Kaoma. The former being urban and the later being rural as such, the differentials in cultural values that shape behaviour in the two areas could explain this finding. Rural women may also be more resilient in copying with food insecurity compared with urban women, as such they may not resort to high-risk sexual behaviour as a copying mechanism in the face of increased vulnerability. Further, the results show that socioeconomic factors are stronger predictors of high-risk behaviour. Thus, food insecurity alone cannot explain high-risk sexual behaviour.

Our findings show that high-risk sexual behaviour is associated with factors at individual, household and community level. Thus our findings confirm the assertions of the socio-ecological model that sexual behaviour is associated with a hierarchy of factors [43]. As Uchidi et al. [35] observed, behavioural interventions that attempt to decrease the odds of risky sexual behaviour by solely focusing on individual behaviours are unlikely to succeed over an extended period of time because the social forces that make women vulnerable to transactional and cross-generational sex are embedded within the sexual norms that are present in the societal conditions in which people live.

## Limitations

The study had some limitations because of the proposed methodology. Firstly, the use a non-experimental design provided a limitation as only associations can be established and not causality. Secondly, self-reported data on food insecurity and high-risk sexual behaviour may be subject to bias. Social desirability and anticipated stigma may influence the reporting of high-risk sexual behaviour. In addition, measurement of HRSB based on self-reporting by one partner may be subject to underestimation of risk to STI and HIV infection as having a high-risk partner may also predispose women to STI and HIV infection. Another limitation is related to the reference period for reporting high-risk sexual behaviour among women aged 18-49. The use of 12 months reference period may be limiting since it does not collect information about sexual history. Conversely, reporting of high-risk sexual behaviour and food-insecurity using the 12 months reference period may be subject to potential recall bias.

## Conclusion

This study found a high prevalence of food insecurity and high-risk sexual behaviour among women of reproductive age. We found that food insecurity is associated with high-risk sexual behaviour before controlling for other variables. We found high odds of engaging in high-risk sexual behaviour among adolescent girls, never married women, women with primary education, women from poor wealth tertile and women in urban areas. Individual level factors alone, cannot explain high-risk sexual behaviour. High risk sexual behaviour is determined by several factors at different levels. Greater attention should be paid to adolescent girls to reduce high-risk sexual behaviour and avert possible consequences. Improving the social-economic status of women through education and economic empowerment among other interventions may reduce vulnerability to HIV infection, unintended pregnancy, improve the health and well-being and ensure that women are not left behind. Further research is required to investigate high-risk sexual behaviour among women beyond the individual level factors. Intervention studies may be useful in identifying possible solutions to reduce high-risk sexual behaviour and avert its consequences among women.

## Data Availability

Data for this study will be made available upon official request to the author. The data provided will be deidentified to protect confidentiality and privacy of study participants.

## Availability of Data and Materials

Data for this study will be made available on request to the corresponding author.

